# Exploring the ‘train the trainer’ model for delivering Making Every Contact Count (MECC) training at scale: A qualitative study

**DOI:** 10.64898/2025.12.11.25342056

**Authors:** Beth Nichol, Angela M. Rodrigues, Sarah Audsley, Anna Haste, Mei Yee Tang, Craig Robson, Jill Harland, Catherine Haighton

## Abstract

**Objectives:** To explore the delivery of the train the trainer (TtT) model for Making Every Contact Count (MECC) training in the North East and North Cumbria (NENC) region of England.

**Design:** A qualitative study, utilising semi-structured interviews.

**Methods:** Interviews were conducted with 21 participants, including MECC TtT trainees (n = 13), eligible non-trainees (n = 6), and principal trainers (n = 2). Data analysis utilised the Framework Method guided by the Theoretical Domains Framework (TDF), and meta-themes were generated that transcended individual TDF domains.

**Results:** Four meta-themes were identified; the need for psychological preparedness to deliver MECC training, successful cascade is influenced by the MECC training content, limited accessibility of the pedagogical approach to both MECC TtT and MECC training, and a need to shift the focus from quantity to quality of MECC training cascade.

**Conclusions:** The TtT model potentially provides unique value for delivering high quality MECC training at scale, providing trainers feel ownership over and able to deliver MECC training. A skills-based approach to MECC training and an experiential learning approach to MECC TtT training is recommended. The MECC TtT programme should provide clear expectations of cascade at sign up stage, allow trainers to adapt content, and evaluate success based on quality.

**Data availability statement:** Fully anonymised transcripts were uploaded onto the UK data service, publicly accessible here: https://reshare.ukdataservice.ac.uk/857461/. The pre-registered protocol is available on Open Science Framework, publicly accessible here: https://osf.io/xz8au.

## Introduction

Making Every Contact Count is a person-centred approach to behaviour change conversations to promote health and wellbeing (1). More evidence is needed to demonstrate a clear effectiveness of MECC on improvements in health and wellbeing of service users (2-4), although evidence is promising to suggest that MECC is acceptable amongst service providers (5-7) and users (8, 9). MECC also compliments the aims of key policy documents in the UK (10-13) and internationally (14) by contributing to a wider culture change towards health promotion and disease prevention (4, 15-17). For example, as part of a holistic and person-centred approach to population health as endorsed by the World Health Organization (WHO) (14), individuals are recognised as active agents with the ability to make choices relating to their own health, including engaging in positive choices described as self-care (18). Subsequently, the WHO recognises the importance of self-care in health promotion and disease prevention on a global scales (18). A consistent message of health promotion through empowerment to engage in self-care behaviours across sectors, for example by applying MECC, may offer the highest probability of health behaviour change and ultimately a reduction in non-communicable diseases on a population level.

MECC was initially proposed, by NHS England (19) within a consensus statement, as an approach to mitigate the rise in non-communicable diseases including cardiovascular disease and cancers through discussing the drivers smoking, alcohol consumption, lack of physical activity, and poor diet (20). More recently, MECC has been applied to behaviour change more holistically to include a wide range of behaviours relevant to the individual (7, 21), in recognition of the wide scope of determinants of health and wellbeing (22). Indeed, a recent consensus study of 40 experts in MECC confirmed that MECC is not defined by the target behaviour, topic of conversation, or even who it is delivered by and where, and instead is defined by its person centred and motivational approach (1). Therefore, it is conceivable that the potential scope and relevance of MECC is far reaching, beyond healthcare where it was initially implemented. However, disseminating the relevant skills and knowledge required to deliver MECC across multiple sectors presents a challenge due to limited resources (15).

The need for a culture change towards prevention and self-care combined with the wide potential scope of MECC has created a demand for the delivery of MECC training at scale. Subsequently, although regional approaches to the delivery of MECC training vary (15, 23, 24), many adopt a ‘Train the Trainer’ (TtT) model to deliver their MECC training (15, 23, 25). The TtT model involves using a principal trainer to provide training to individuals so that they become MECC trainers, cascading MECC training to those they go on to deliver training to. When successful, the TtT model thus offers a wider potential reach of training with the same amount of resources (26, 27). Additionally, particularly given that the wide scope of MECC means that individuals from any role or setting could receive MECC training, the TtT model means that MECC trainers are more likely to possess specific knowledge of their audience, increasing its perceived relevance (26, 27).

Despite the increasing utilisation of the TtT model to deliver MECC training (15, 25), there lacks a qualitative evaluation to better understand the barriers and facilitators to implementing the TtT model. Given the early stage of research, qualitative analysis is appropriate to explore the experiences of individuals who have received MECC TtT training, the challenges they faced in cascading training, and their thoughts on the suitability of the TtT model for the delivery of MECC training. Thus, this study reports the in-depth qualitative analysis of the second stage of a mixed methods sequential study that evaluated the TtT model of MECC training across the North East and North Cumbria (NENC) region of England, reported elsewhere (28). The aim of this stage was to explore the in-depth explanations for the low rate of cascade of MECC training identified in stage one.

## Methods

### Study design and setting

The current study describes the themes generated from a qualitative analysis of interview and survey data collected as part of the wider study (28). The study adopted a pragmatic approach, applying behavioural science to provide a framework for barriers and enablers to training cascade. Namely, the Theoretical Domains Framework (TDF) was applied which divides behavioural determinants into 14 domains corresponding to capability, opportunity, or motivation to perform a specified behaviour (29). Reporting of the following methods adhered to the Consolidated criteria for reporting qualitative research (COREQ, see Supplementary Material 1) (30), with the caveat that information power was selected as a more appropriate and valid sample size justification than data saturation (see further explanation below) (31). As part of the wider mixed methods study, a professional contributor panel of four contributors was formed to advise on the study (more details are provided in the mixed methods paper: (28)). Prior to data collection, the study protocol for the full mixed methods study was preregistered via Open Science Framework (https://osf.io/xz8au).

This study was approved by the Ethics department at Northumbria University (Project ID 5033). This study also received R&D approval from the North East and North Cumbria Integrated Care System (ICS) as a service evaluation, as determined by the Health Research Authority.

### Participants and recruitment

To explore potential barriers to accessing MECC TtT training, interview participants (n = 21) were eligible for MECC TtT training but may not have received it. Participants were adults (>18) residing in the UK. Sampling was purposive to ensure that the sample was representative of current implementation of the TtT programme in terms of sectors (e.g healthcare, local authority, third sector), level of training delivery or uptake (including those who had attended and cascaded training and those who had not), and type of participation in training (self-referred or delegated to attend). The sample was organised into three distinct groups; principal trainers (n = 2), those who completed the MECC TtT programme (n = 13), and those who were eligible but had not completed the TtT programme (n = 6). Of those who had completed the programme, two participants were also involved in the implementation of MECC TtT and thus were also able to provide insights on the success of others in becoming trainers.

The Regional Delivering MECC at Scale Coordinator for the NENC and the MECC at Scale TtT course trainer (principal trainer) were asked to provide eligible participants with the study information sheet and expression of interest form which potential participants could complete via a Qualtrics (32) link (32). The Regional Delivering MECC at Scale Coordinator utilised existing networks and contacted potential participants via email, within online Teams meetings (e.g MECC strategy group), and in-person (e.g at TtT sessions). When recruitment occurred in person, expression of interest forms were available through paper copies. Recruitment also took place via alternative routes such as third sector networks and existing individual contacts using the same approach. To facilitate purposive sampling, expression of interest forms collected optional basic demographic data (occupation, setting, and receipt of MECC training). In addition, the principal trainer participated in an interview. Thus, recruitment occurred on an opt-in basis.

Sample size for qualitative interviews was defined apriori using the model of information power by Malterud, Siersma and Guassora (31). Target sample size was estimated from the aim, specificity of sample, use of theory, interviews, and analysis strategy. As the aim was relatively specific, sampling was purposive, analysis used an established theoretical framework and took a pragmatic approach, and the primary researcher (BN) was experienced in qualitative research and aimed to build a rapport with participants before interviewing, the estimated total required sample size was around 20.

Additionally, as part of a survey collected pre and post attendance to MECC TtT training, responses to the seven free text questions (see Supplementary Material 2) included in the post survey evaluation (n = 373), whcih achieved a 72% response rate of all attendees across the NENC between May 2022 to 2^nd^ November 2023 (point of retrieval), were included in the qualitative analysis to increase generalisability of the findings (more details of the post training survey is provided in the mixed methods paper: (28)).

### Materials and data collection

Two topic guides were developed, tailored to individuals who had or had not attended MECC TtT training. Topic guides were informed using the TDF (29, 33) and pilot tested and further refined after discussion with the professional contributor panel. For principal trainers (one delivered MECC TtT not as part of the regional offer), the topic guide was applied more flexibly to reflect their different role and insight in MECC training cascade. Topic guides (see Supplementary Material 2) explored the barriers and facilitators to training cascade, any existing strategies or suggestions to improve training cascade, experience of and reflections on the TtT programme and associated resources (e.g bi-monthly trainer forum), and thoughts on the TtT model of training delivery. For those who had not attended MECC TtT training, the topic guide focused more on their motivations to attend and barriers or facilitators to accessing MECC TtT training. Additionally, free text post survey questions (Supplementary Material 3) centred around the training experience, whilst the interviews also asked about accessing the training and experiences in attempting to cascade MECC training after the TtT training.

After potential participants expressed interest to take part, a time and location to conduct an interview was arranged. Participants who were outside of healthcare were provided with the option of an in person or online interview via Teams, and interviews with participants within healthcare were conducted online. Thus, three interviews were conducted in person and the remaining 18 online. The primary researcher (BN) that conducted all semi-structured interviews is female and experienced in both conducting interviews and with MECC; including previous attendance to the MECC TtT programme. Thus, the interviewer had previously met and built rapport with some participants, and built rapport with any participants where a relationship had not previously been established. The interviewer introduced themselves to all participants as a research assistant on the project. One-to-one interviews flexibly applied one of the two topic guides (described above). Interviews lasted between 18 and 88 minutes and were digitally recorded and transcribed verbatim via a transcription service and anonymised.

### Data analysis

Qualitative data (interviews and free text comments) was analysed using the Framework Method (34). To facilitate familiarisation with the data, all interviews were conducted and analysed by the primary researcher (BN) through line-by-line coding. Namely, themes were formed deductively using the TDF domains (29), however within each domain inductive analysis was applied whereby codes were built into sub-themes. Interview data and post survey comments were analysed separately although analysis of the free text comments was used to inform sub themes identified using the interview data. Given the lack of context provided within the free text comments, their analysis was supplementary to the interview data and added to existing themes formed from the interview data. A 10% sample of survey and interview data was independently coded to ensure inter rater reliability of TDF coding (by SA and MYT, respectively), with disagreements resolved through discussion between all three independent raters (BN, MYT, and SA). Additionally, as conducted within previous research (35), meta-themes were developed by the primary researcher (BN) accompanied by discussion with the core research team (CH and AMR). Meta-themes transcended single TDF domains and were inductively created, although informed by sub-themes and codes within each domain.

A coding system was used to label transcripts of principal trainers (PT), those who had attended the MECC TtT training (AT), and those who had not attended the MECC TtT training (NA), respectively. In accordance with open data procedures, fully anonymised transcripts were uploaded onto the UK data service (36), a public repository, with the consent of participants using an ‘as open as possible, as closed as necessary’ principle (37). Namely, given that the nature of the topic was not overly sensitive or personal for participants it was judged appropriate to upload transcripts onto a public repository provided informed consent was attained. All data analysis was conducted via Nvivo 12 Pro (38).

## Results

### Themes and sub-themes guided by the TDF

Specific barriers and facilitators according to TDF domains are described in the mixed methods paper (28). The main sub-themes within each TDF domain and corresponding example codes are provided in Table 1 and the full coding framework (including example quotes) is provided in Supplementary Material 3. Participants placed value on individuals who were interested in MECC self-selecting themselves to become MECC trainers as opposed to mandatory training with disinterested attendees (Intentions). However, top-down drivers were perceived to be much more valuable. For example, a reported difficulty in identifying an audience was most often attributable to a limited staff capacity to attend MECC training (a top-down influence) rather than a lack of interest in MECC (a bottom-up influence). Thus, the paucity of incentives to deliver MECC training meant that benefits for MECC trainers to delivering MECC training without top-down support was low (Reinforcement). Also, some participants suggested that ‘TtT’ is not clear on what it requires from attendees (Environmental Context and Resources), demonstrable by the finding that many attendees were unaware that they were attending a TtT programme (Knowledge) and instead wanted to learn more about MECC rather than deliver MECC training (Goals).

**Table 1:**
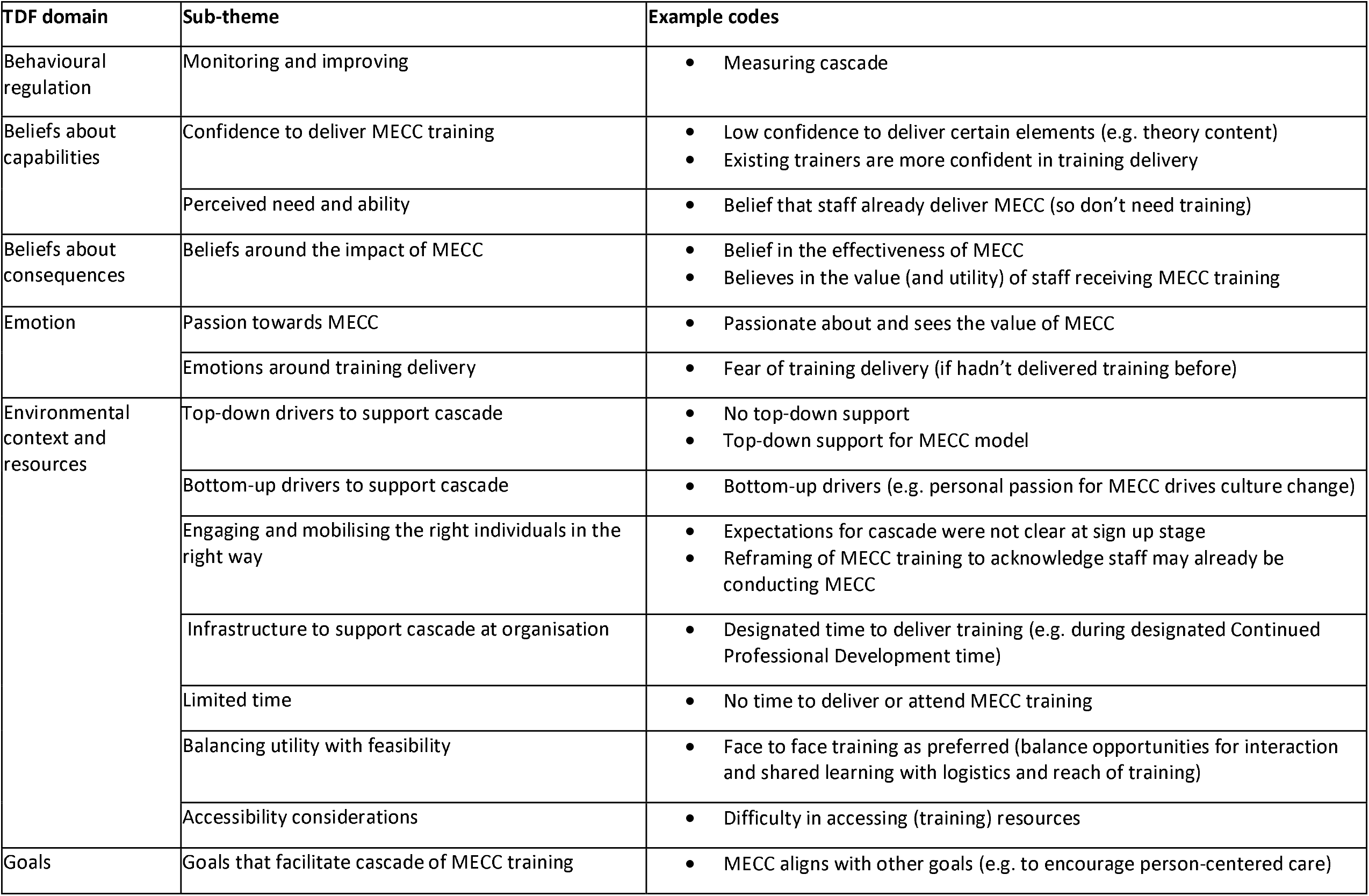
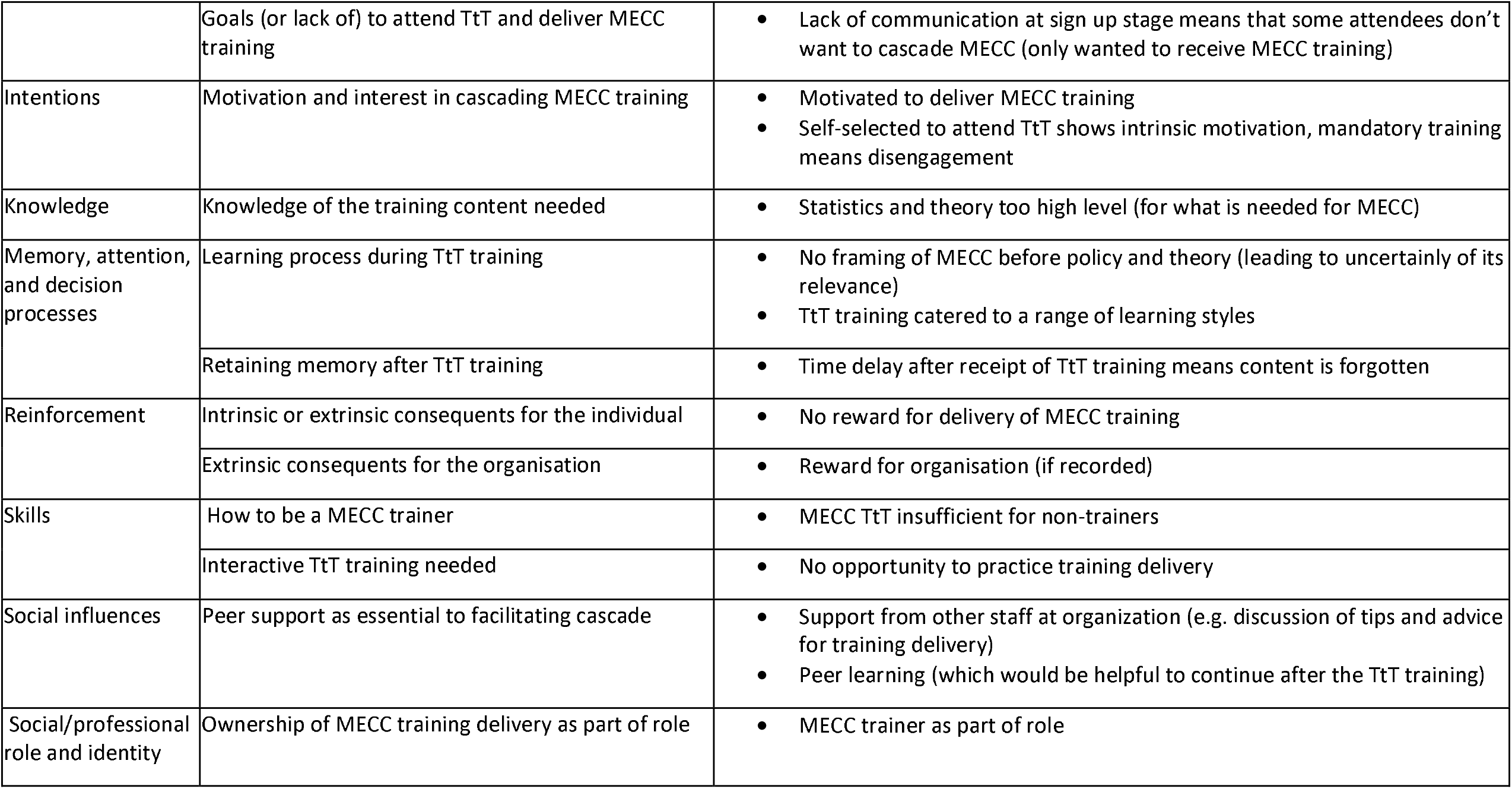
Main sub-themes within each domain and example corresponding codes from the overall coding framework Supplementary Material 1. The TDF domain optimism is not included as its relevance to the data was minimal.

| TDF domain | Sub-theme | Example codes |
| --- | --- | --- |
| Behavioural regulation | Monitoring and improving | Measuring cascade |
| Beliefs about capabilities | Confidence to deliver MECC training | Low confidence to deliver certain elements (e.g. theory content)<br>Existing trainers are more confident in training delivery |
|  | Perceived need and ability | Belief that staff already deliver MECC (so don't need training) |
| Beliefs about consequences | Beliefs around the impact of MECC | Belief in the effectiveness of MECC<br>Believes in the value (and utility) of staff receiving MECC training |
| Emotion | Passion towards MECC | Passionate about and sees the value of MECC |
|  | Emotions around training delivery | Fear of training delivery (if hadn't delivered training before) |
| Environmental context and resources | Top-down drivers to support cascade | No top-down support<br>Top-down support for MECC model |
|  | Bottom-up drivers to support cascade | Bottom-up drivers (e.g. personal passion for MECC drives culture change) |
|  | Engaging and mobilising the right individuals in the right way | Expectations for cascade were not clear at sign up stage<br>Reframing of MECC training to acknowledge staff may already be conducting MECC |
|  | Infrastructure to support cascade at organisation | Designated time to deliver training (e.g. during designated Continued Professional Development time) |
|  | Limited time | No time to deliver or attend MECC training |
|  | Balancing utility with feasibility | Face to face training as preferred (balance opportunities for interaction and shared learning with logistics and reach of training) |
|  | Accessibility considerations | Difficulty in accessing (training) resources |
| Goals | Goals that facilitate cascade of MECC training | MECC aligns with other goals (e.g. to encourage person-centered care) |
|  | Goals (or lack of) to attend TtT and deliver MECC training | Lack of communication at sign up stage means that some attendees don't want to cascade MECC (only wanted to receive MECC training) |
| Intentions | Motivation and interest in cascading MECC training | Motivated to deliver MECC training<br>Self-selected to attend TtT shows intrinsic motivation, mandatory training means disengagement |
| Knowledge | Knowledge of the training content needed | Statistics and theory too high level (for what is needed for MECC) |
| Memory, attention, and decision processes | Learning process during TtT training | No framing of MECC before policy and theory (leading to uncertainty of its relevance)<br>TtT training catered to a range of learning styles |
|  | Retaining memory after TtT training | Time delay after receipt of TtT training means content is forgotten |
| Reinforcement | Intrinsic or extrinsic consequents for the individual | No reward for delivery of MECC training |
|  | Extrinsic consequents for the organisation | Reward for organisation (if recorded) |
| Skills | How to be a MECC trainer | MECC TtT insufficient for non-trainers |
|  | Interactive TtT training needed | No opportunity to practice training delivery |
| Social influences | Peer support as essential to facilitating cascade | Support from other staff at organization (e.g. discussion of tips and advice for training delivery)<br>Peer learning (which would be helpful to continue after the TtT training) |
| Social/professional role and identity | Ownership of MECC training delivery as part of role | MECC trainer as part of role |

On attending the MECC TtT programme, a high level of knowledge was required to deliver the MECC training content around public health statistics, health inequalities, and theory (Knowledge), thus participants commonly felt a lack of confidence to deliver the content (Beliefs about Capabilities). Participants were unclear on how much of the complex information related to MECC delivery so found it difficult to consolidate it (Memory, Attention, and Decision-Making Processes) especially if they had no experience in training delivery, which created stress or discomfort around delivering MECC training (Emotions). Face-to-face training encouraged facilitators such as shared learning and interaction but the logistical challenges of face-to-face training were acknowledged by participants (Environmental Context and Resources). Refresher training was thought to update knowledge, aid memory of content, revive motivation to deliver training, and encourage social support (Environmental Context and Resources).

After receipt of MECC TtT training, some participants discussed their concern that staff in their organisation were already competent in the skills required to deliver MECC and engaged in MECC conversations without the need for training (Beliefs about Capabilities). Subsequently, these participants expressed uncertainty of how to approach MECC training in order to acknowledge the existing competency of the audience. Also, a time delay between attending the MECC TtT programme and cascading MECC training made cascading less likely as momentum was lost (Environmental Context and Resources) and content was forgotten (Memory, Attention, and Decision Making Processes).

### Meta-themes that transcend TDF domains

Four meta-themes were identified (see Table 2 for example quotes). Each meta-theme is discussed in turn.

**Table 2:**
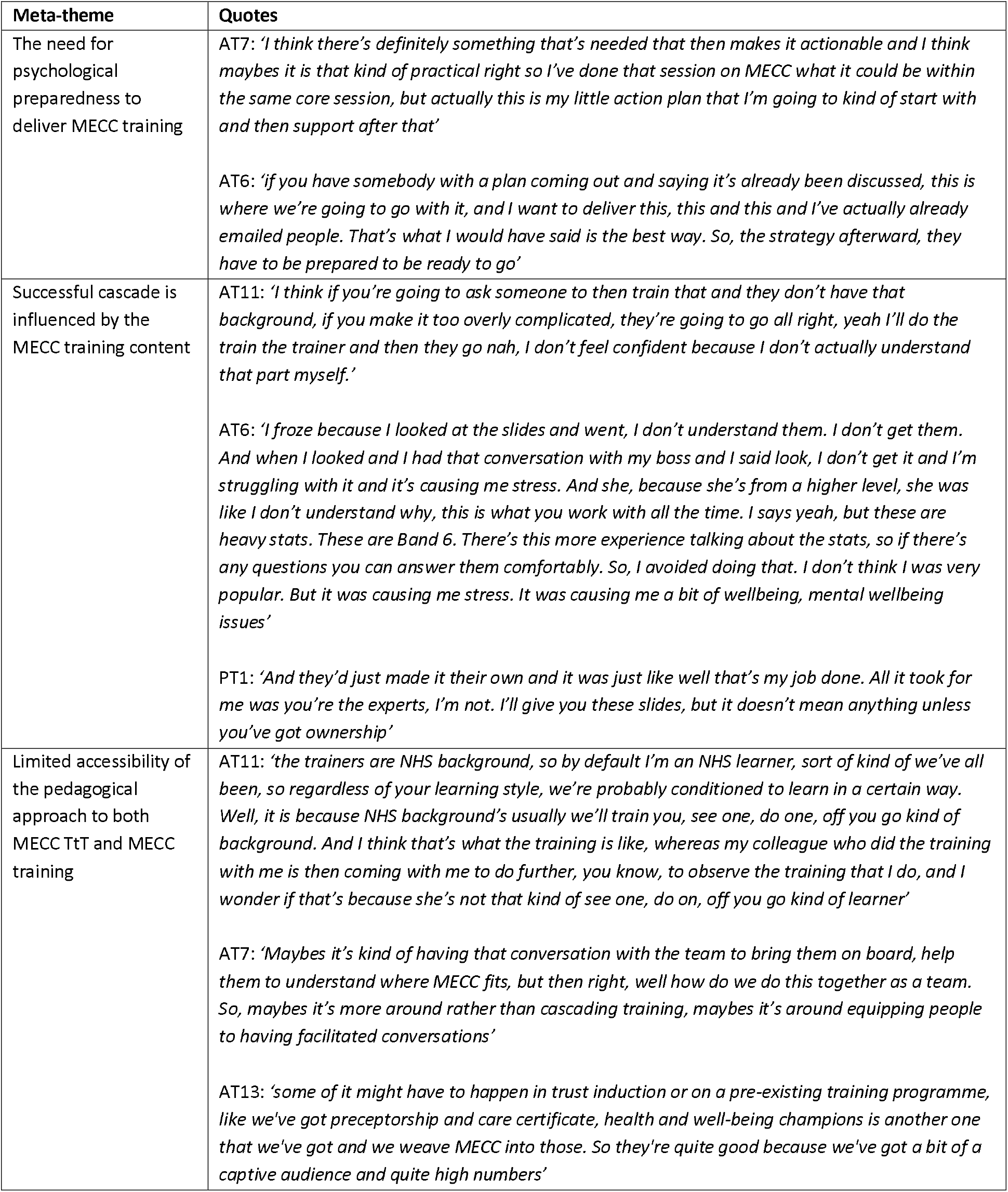
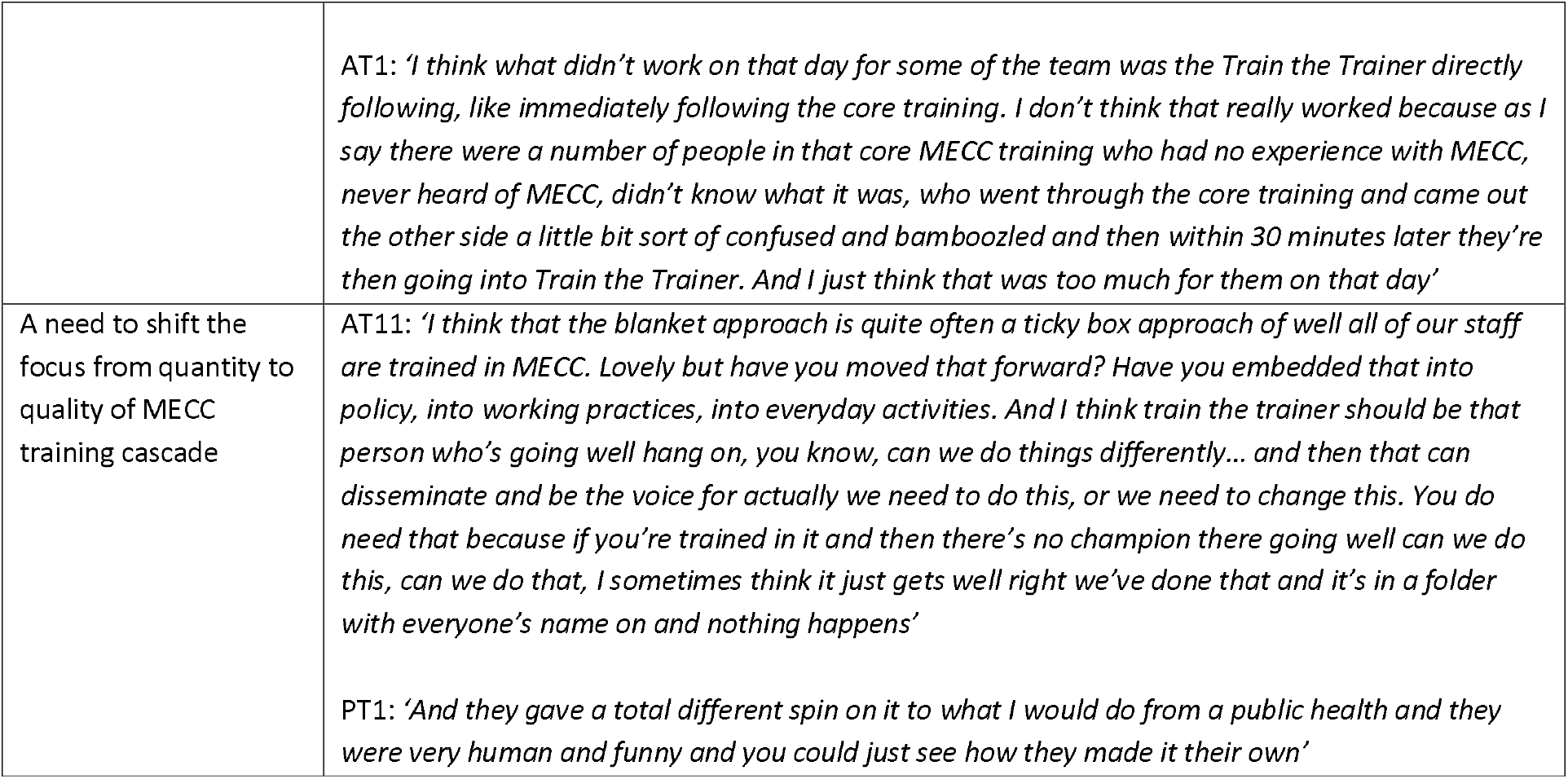
Corresponding quotes for each of the four meta-themes.

#### The need for psychological preparedness to deliver MECC training

From the post survey comments it was apparent that many participants showed no intention of utilising the MECC TtT programme to deliver training, with some noting their surprise at the TtT training. Many TtT attendees were not psychologically prepared to become a MECC trainer. Yet as the MECC TtT training was readily available, individuals were nonetheless encouraged to complete it. Individuals felt more ready to deliver MECC training if it helped to fulfil their role and achieve strategic goals that were set by leadership and management. Active involvement in the planning and goal setting for MECC training delivery created more motivation and preparedness to deliver MECC training than if goals were imposed onto them.

#### Successful cascade is influenced by the MECC training content

Relatedly, active involvement in tailoring the MECC training content encouraged personal ownership over the content which facilitated personal responsibility to cascade, which was particularly helpful for trainers whom delivery of MECC training was not part of their role. Furthermore, ownership over the course content encouraged training cascade.

MECC training content could also in itself be a barrier to cascade. The high informational load of the course content meant that participants reported feelings of overwhelm and required a lot of time to familiarise themselves with it after the MECC TtT programme. Specifically, those who were newly exposed to the statistics and theory content proposed that it was overcomplicating what they were being asked to do. Also, some participants felt limited by their abilities to amend the content accordingly for worries of compromising on training quality and consistency.

#### Limited accessibility of the pedagogical approach to both MECC TtT and MECC training

Despite participants reporting that a range of learning styles were catered to within the training, a didactic training approach meant that some participants reported feelings of overwhelm at immediately learning how to deliver MECC training after receiving it themselves. Whilst participants from healthcare settings struggled acutely with time and capacity issues, participants within these settings tended to be familiar with the didactic training style and TtT model of training delivery, particularly those with nursing backgrounds. Thus, combined with possessing higher relevant knowledge relating to MECC and feeling MECC to be part of their role, participants within healthcare settings were more confident with the pedagogical approach of the MECC TtT programme whilst participants from other settings required more support.

Considering the learning approach of the MECC training, a minority of participants discussed alternative approaches to dissemination of MECC principles. For example, informal training delivery through conversations with other staff during existing meetings was considered to be more feasible and to encourage a culture change. Others shortened MECC training to make it more accessible or delivered it as part of other training programmes in order to frame MECC. Such seemingly beneficial adaptations suggested a need to consider the definition of ‘successful’ cascade of MECC training.

#### A need to shift the focus from quantity to quality of MECC training cascade

Monitoring was mainly discussed in relation to achieving organisational level targets concerning MECC delivery, through numbers of individuals who had received TtT training, number of MECC training sessions delivered, and number of individuals delivered to. Whilst easily estimating the success of the TtT model through number of individuals trained could be motivating, it could also encourage undue focus on numbers rather than quality of TtT training and the value of the subsequent cascaded MECC training. Combined with a wide availability of the MECC TtT training, top-down support often encouraged attendance at the MECC TtT training but did not extend further than attendance, including the preparation, planning, and time and audience needed to cascade MECC training. Thus, there was an overall focus on individuals completing the MECC TtT training, although this was insufficient to encourage cascade.

Despite this, participants recognised that there was a unique value to the TtT model in that it encouraged trainers with knowledge of the organisations and individuals they were delivering training to, thus increasing the quality of MECC training that was delivered.

## Discussion

In summary, a lack of communication about the programme, a TtT training programme that is more readily available than MECC training itself, and top-down support to attend the TtT training but not cascade MECC training means that most individuals who attend the TtT training across the NENC are not ready or willing to deliver MECC training. Furthermore, the MECC TtT programme across the NENC is relatively inflexible and inaccessible for individuals outside of healthcare settings and those with limited existing public health knowledge and training experience. Also, a didactic TtT training style does little to encourage ownership of MECC training for MECC trainers, which is essential in maintaining motivation to deliver training particularly when there are no incentives or expectations to cascade training. Whilst ownership of the MECC training content could be facilitated through opportunities for MECC trainers to tailor the MECC training, this flexibility was hampered by concerns of compromising the quality of MECC training and there was a lack of guidance in navigating such worries. Ironically, a focus on delivering the TtT programme as intended by monitoring ‘success’ through attendees of the TtT training and cascade rates overlooked the added value of the TtT model in encouraging MECC training that was relevant to its audience. Finally, for bottom-up processes to become more impactful in encouraging MECC training delivery, MECC TtT training must focus on the ‘why’ and ‘how’ rather than provision of information, or the ‘what’, alone. Many of the challenges surrounded a balance between the flexibility of MECC training (which was a strength) and the extent to which MECC training can be amended to retain its quality and consistency. However, personal ownership and buy-in of the training content is a key factor for successful cascade (39-41), which can be achieved through modifying the content and the teaching style and trainers adding their own experiences (39, 41, 42). The ability to amend training content in turn also increases confidence to deliver training (39). Furthermore, ownership and buy-in to the training has been found to be accelerated through tacit knowledge (39) or experiential learning (40). In contrast to the mostly didactic approach to the MECC TtT programme reported by participants, experiential learning encourages trainees to actively engage in their own learning, including through practical experience and reflection (43). For example, the healthy conversation skills (HCS) approach to MECC training, focuses less on imparting public health knowledge and more on teaching trainees the skills to conduct motivational conversations (44), and has its own approach to the TtT model that mirrors the reflective HCS principles (25). Furthermore, HCS training has been repeatedly demonstrated to encourage service providers to deliver MECC (6, 9, 45-47). Consequently, the HCS TtT approach disseminates effective training content which could facilitate successful cascade itself, encouraged through an experiential learning TtT approach. Thus, the themes identified within this study appear to be tightly interconnected and support a HCS approach to delivering MECC training through the TtT model. Through facilitating buy-in and ownership over MECC training, individuals come to see themselves as ‘MECC trainers’ (39). Findings from the current study indicate that the content of training delivered is inherently predictive of the success of the TtT model that aims to disseminate it However, it is important to note that beyond the characteristics of the TtT training itself, organisational level support is key to determine successful cascade (25).

Beyond its potential for greater efficiency, the TtT programme does appear to facilitate the delivery of MECC training that is relevant to the individuals it is delivered to and this asset should be fostered and encouraged. Namely, trainers with knowledge of the local area and organisation could provide context to MECC that may help to increase the quality of MECC training thus its outcomes including satisfaction of trainees and behaviour change. Indeed, in addition to cost-effectiveness (26), specific benefits of the TtT model include local knowledge (26) and encouragement of collaboration across sectors (48), with many participants within the current study citing face to face training as an opportunity to network. Thus, there remains rationale to optimise the TtT model rather than abandon it.

### Strengths and limitations

A key strength of this study was the holistic perspective of the inclusion of those who had received MECC TtT training, were eligible but not received it, and principal trainers, individuals from across healthcare, local authority, and voluntary and community settings, and those who had and had not gone on to cascade MECC training. Subsequently, this created a representative sample that allowed for a comprehensive exploration into the TtT model to deliver MECC training. However, a limitation of the current study is that it focused on the NENC approach to both MECC training and the TtT programme which may limit the generalisability of the findings. Indeed, regional approaches can vary vastly including HCS (44). Whilst the findings from the current study can inform on the challenges of TtT models and the factors that facilitate successful delivery more generally including to deliver MECC training, future research would benefit from directly comparing different MECC training and TtT approaches to the delivery of MECC training, as the current study found the MECC training content to be an important barrier to successful cascade as well as the didactic TtT approach.

### Implications for practice

To facilitate successful cascade of high quality MECC training, action is needed on programme, individual, and organisational levels. On a programme level, it would be helpful to reconsider the pedagogical approach to make it more accessible to non-healthcare settings, including exploring more informal ways of disseminating MECC principles. It would also be helpful to review the reliance on theory and statistics within the MECC training and consider a more reflective, skills-based approach such as HCS. On an individual level, MECC trainers should be allowed flexibility to tailor and take ownership over the MECC training content and engage in tacit and experiential learning related to how to cascade the MECC training to others. On an organisational level, top-down ownership is needed from respective organisations that extends beyond just TtT attendance to enabling actual cascade through provision of time and resources, as is a shift of focus from quantity to quality of cascade by measuring impact rather than just numbers trained. With these changes, MECC trainers’ local knowledge can be utilised to make MECC training more relevant and impactful.

## Conclusion

In conclusion, many factors influence perceived ownership over MECC content, which in turn facilitates MECC trainers who perceive MECC training delivery as part of their identity. The content of the MECC programme is vital for determining cascade, as is the pedagogical approach to facilitate the cascade of the content. Furthermore, more value should be placed on the quality of cascade and how this can best be achieved rather than the ‘*success*’ of cascade through statistics alone, to achieve the primary aim of the TtT to deliver MECC at scale.

## Data Availability

All data produced are available online at https://reshare.ukdataservice.ac.uk/857461/

## Data Availability

All data produced are available online at https://reshare.ukdataservice.ac.uk/857461/

