## Supplementary Material for "Exploring the ‘train the trainer’ model for delivering Making Every Contact Count (MECC) training at scale: A qualitative study"

**Supplementary Material 1:** Topic guides for a) non-completers of MECC TtT training and b) completers of MECC TtT training.

**Supplementary Material 1a)**

Briefly re-iterate the study:

- Check they have read the information sheet and give consent

My name is Beth Nichol, and I am research assistant on a project exploring how to optimise the train the trainer model to deliver MECC training at scale. As part of this, we are interviewing both those who have been invited and not attended, and those who have attended the train the trainer MECC training. We want to explore your experiences to look at how the training can be improved.

In case you are not familiar with MECC, it stands for Making Every Contact Count, which is a public health initiative that aims to make use of the thousands of interactions service providers have with service users or patients every day, to talk about health and wellbeing topics such as smoking, alcohol, diet, and physical activity. Whilst MECC began within healthcare, recently training has expanded to the VCSE, police and fire services, and beyond. The core MECC train the trainer training delivers the core principles of MECC, with the aim that trainees go on to deliver MECC training to frontline staff within their organisation.

If you don’t mind, the interview will be recorded and anonymously transcribed and I will take some notes. The notes and recording will be kept completely private, meaning no names or identifiable information that you mention in the recording will be used when it is typed up. This means that the transcript will not identify you in any way. Having said that, you may want to stick to first names and avoid using identifiable information for the sake of the recording, but this is entirely your choice. Are you happy to go ahead with the discussion?

**Part 1: Existing awareness of MECC**

Can you first describe a little bit about your organisation and your role within it?

- What motivated you to work in this role?
- What are your thought regarding the suitability of health promotion within your organisation?
- Can you describe any other activities around health promotion that you are engaged with as part of your role within your organisation?

Can you recall how you first became familiar with the term MECC?

- Can you describe any awareness of MECC you had at the point of being invited to complete MECC training?
- Can you describe how you feel about MECC as an approach?
- How would you describe your understanding of MECC?
- How would you describe your motivations to deliver MECC training to others?
- How would you describe your motivation to deliver MECC conversations?
- Can you describe any instances where you have delivered a MECC conversation?
- Have you accessed any other MECC training packages before? If so, can you describe these?
- Can you describe any other training you have had that provided you with skills that facilitated MECC?

**Part 2: Accessing the training**

Can you describe what the organisational culture is surrounding MECC in your organistion? (e.g awareness and encouragement of MECC)

Were you invited to complete MECC TtT training? If so, can you describe how you were invited?

- Why do you think you were invited to participate in MECC train the trainer training?
- What were your thoughts regarding your suitability for receiving the TtT training compared to other people from your team/organisation?
- Can you remember what your superiors (if applicable) thought about you attending MECC TtT training?
- How was the training offered to you (e.g online, face to face)? Did this affect your decision to reject or accept the training?
- What were your thoughts in terms of accessibility of the MECC TtT training (e.g date, time, venue)?
- Were you aware of the resources available to help you deliver MECC training after receiving TtT training?
- Does your organisation have a MECC implementation plan? Does this affect your intentions in any way?

**Part 3: Barriers and facilitators to MECC TtT training**

Can you describe why you did not attend the MECC train the trainer training?

- Was there anything that prevented you from attending the train the trainer training?
- Can you describe anything that might have helped you to attend the train the trainer training?
- Do you have any intentions to complete MECC TtT training in the future?

Do you have any thoughts regarding the train the trainer model?

- What are the barriers to this model?
- Do you think there are any strengths to this model?
- Do you have any thoughts regarding whether the TtT model is best for delivering MECC at scale?
- Do you have any thoughts regarding how helpful the TtT model is for ultimately encouraging MECC conversations with the end user?

**Part 4: Think aloud**

What comes to mind about the following strategies for improving training cascade?:

- Provision of refresher MECC training sessions for trainers
- Provision of peer support networks or groups where resources, experiences, and knowledge could be shared

**Part 5: Further comments**

Is there anything else you would like to add?

**Supplementary Material 1b)**

Briefly re-iterate the study:

- Check they have read the information sheet and give consent

My name is Beth Nichol, and I am research assistant on a project exploring how to optimise the train the trainer model to deliver MECC training at scale. As part of this, we are interviewing both those who have been invited and not attended, and those who have attended the train the trainer MECC training. We want to explore your experiences to look at how the training can be improved.

In case it would be helpful to be provided with a refresher of what MECC is, it stands for Making Every Contact Count, which is a public health initiative that aims to make use of the thousands of interactions service providers have with service users or patients every day, to talk about health and wellbeing topics such as smoking, alcohol, diet, and physical activity. Whilst MECC began within healthcare, recently training has expanded to the VCSE, police and fire services, and beyond. The core MECC train the trainer training delivers the core principles of MECC, with the aim that trainees go on to deliver MECC training to frontline staff within their organisation.

If you don’t mind, the interview will be recorded and anonymously transcribed and I will take some notes. The notes and recording will be kept completely private, meaning no names or identifiable information that you mention in the recording will be used when it is typed up. This means that the transcript will not identify you in any way. Having said that, you may want to stick to first names and avoid using identifiable information for the sake of the recording, but this is entirely your choice. Are you happy to go ahead with the discussion?

**Part 1: Existing awareness of MECC**

Can you first describe a little bit about your organisation and your role within it?

- What motivated you to work in this role?
- What are your thoughts regarding the suitability of health promotion within your organisation?
- Can you describe any other activities around health promotion that you are engaged with as part of your role within your organisation? (e.g smoking, weight management)
- Can you tell me about the training you have received around MECC? (e.g core MECC)
- Have you accessed any other MECC training packages before? If so, can you describe these?
- Can you describe any other training you have had that provided you with skills that facilitated MECC?

I would like you to think back to before you received MECC training.

- Were you familiar with MECC before receiving TtT training? Can you recall how you first became familiar with the term MECC?
- Can you describe any awareness of MECC you had at the point of being invited to complete MECC training?
- Can you describe how you felt about MECC as an approach, and has this changed over time?
- Can you remember what your understanding was of MECC before the TtT training?
- Can you describe any MECC (or healthy lifestyle) conversations you delivered before receiving training?
- Can you remember what you motivation levels were like to deliver MECC conversations before accessing the TtT training?
- Before receiving the training, can you remember what you motivation levels were like to deliver MECC training to others?

**Part 2: Accessing the training**

Can you describe how you were invited to complete the MECC train the trainer training?

- Why do you think you were invited to participate in MECC train the trainer training?
- Why did you attend MECC TtT training? (nominated, organistional or personal motivations) Did anyone encourage you to attend?
- What motivated you to attend and not decline the training?
- What were your thoughts regarding your suitability for receiving the TtT training compared to other people from your team/organisation?
- Can you describe what the organisational culture is surrounding MECC in your organistion? (e.g awareness and encouragement of MECC, executive support)
- Can you remember what your superiors (if applicable) thought about you attending MECC TtT training?
- How was the training offered to you (e.g online, face to face)? Did this affect your decision to reject or accept the training?
- What were your thoughts in terms of accessibility of the MECC TtT training (e.g date, time, venue)?

**Part 3: Experience of the training**

Can you describe your experience of the MECC TtT training?

- What did you expect from MECC TtT training?
- What did others expect of you on attending MECC TtT training?
- Did the training meet your expectations? Why/why not?
- Was there any aspects of the training you particularly liked? If so, can you describe these?
- Can you describe anything you would improve about the TtT training?
- Do you have any thoughts regarding the duration of the MECC TtT training?
- Did you understand everything that was communicated to you during training?
- What did you think about the level of information that was provided? (too in-depth, too easy, just right)
- How did you find balancing your learning for MECC TtT training with your role?
- Did you have any thoughts regarding the suitability of the training content for your target audience?

Can you describe anything you took away from the TtT training (e.g thoughts, feelings, knowledge)?

- What aspects of the course content affected this motivation?
- Can you describe how you felt about your understanding of MECC after TtT training?
- What aspects of the course affected your understanding of MECC?
- Can you describe any skills you learnt related to delivering MECC training?
- What aspects of the course were particularly useful for developing skills related to delivering MECC training?
- Can you describe what aspects of the course affected your confidence and motivation to deliver MECC training after receiving the TtT training?
- How did you feel in your overall ability to deliver MECC training after the TtT training?
- Do you feel capable of delivering training after recieving the TtT training? Why/ why not?
- Do you have the opportunity to deliver MECC training? Why/ why not?
- Have you ever attended other train the trainer training before? If so, how did the MECC training compare?
- What other ways of learning about delivering MECC training might you have benefitted from? (Probe- What sort of training approach has worked for you in the past)
- How did you find the current approach? (Probe - What other ways of sharing the learning with colleagues do you think might work as an alternative?)

If you were to attend the training again, what would you want the trainer to focus more on and what would you want them to spend less time on?

**Part 4: Training-cascade**

Have you delivered MECC training since receiving Train the trainer training?

- Did you make any plans to deliver MECC training after receiving TtT training?
- Who, if anyone, have you delivered MECC training to? How did you think that went?
- How did you find the process of arranging and delivering training?
- Can you describe anything you’ve changed that differs from the core MECC training you received?
- Can you describe anything that has prevented you from delivering MECC training?
- What challenges have you faced in cascading the training down?
- Is there anything that has particularly helped you deliver MECC training?
- What would your recommendations be to help people like you cascade the training down?

Can you describe any resources you found to be particularly helpful in helping you cascade training? (e.g NHS futures, MECC gateway, MECC link)

- Were you aware of the resources available to help you deliver MECC training after receiving TtT training?
- Can you describe any resources you have had access to that have assisted you with delivering training?
- How did you find the process of accessing the relevant resources?
- Do you have any thoughts regarding the suitability of the resources for your target audience?
- Did you amend the resources in any way? If so, can you describe why and how you amended them?
- Do you have any suggestions of how the resources could be amended to better support your delivery of MECC training?
- Can you describe any resources outside of those provided to you that have helped you deliver MECC training? (including any you created)
- Are there any further resources that you would like to have access that would help you deliver MECC training?

Can you describe any support you found to be particularly helpful in helping you cascade training? (e.g leadership, strategy group)

- Have you received any support after (e.g trainers forum)? If so, please describe what support you have received?
- Can you describe any support networks or specific people that have helped you to deliver MECC training?
- Does your organisation have a MECC implementation plan? Does this affect your intentions in any way?
- Can you describe any other ways that senior leadership/management at your organisation are invested in the delivery of MECC training?

Do you have any thoughts regarding the train the trainer model?

- What are the barriers to this model?
- Do you think there are any strengths to this model?
- Do you have any thoughts regarding whether the TtT model is best for delivering MECC at scale?
- Do you have any thoughts regarding how helpful the TtT model is for ultimately encouraging MECC conversations with the end user?

**Part 4: Think aloud**

What comes to mind about the following strategies for improving training cascade?:

- Provision of refresher MECC training sessions for trainers
- Provision of peer support networks or groups where resources, experiences, and knowledge could be shared

**Part 5: Final comments**

Is there anything else you would like to add?

**Supplementary Material 2: Post survey free text questions**

- Is there anything you would change within the training or any further information that would help?
- What did you find most useful from today’s training?
- What will you change in your practice as a result of this training?
- Is there anything else you would like to tell us about today’s training?
- If you require some support, what support do you require?
- What is the most important thing you got from your MECC training?
- Any comments or suggestions that may help us to improve the training?

**Supplementary Material 3:** Coding framework as guided by the TDF, with sub-themes presented within each domain. Example codes are provided although they are not an exhaustive list. PSC = post-survey comments.

| TDF domain | Sub-theme | Example code | Example quote |
| --- | --- | --- | --- |
| Behavioural regulation | Delivering high quality training | Problem solving | AT7: *‘I think there’s definitely something that’s needed that then makes it actionable and I think maybes it is that kind of practical right so I’ve done that session on MECC what it could be within the same core session, but actually this is my little action plan that I’m going to kind of start with and then support after that. I just think there probably has to be maybes some steps to being able to do that because ultimately, I think that’s how we begin to understand what the sticking points are, what the barriers are, what support is needed’* |
|  | Monitoring and improving | Measuring cascade | NA5: *‘if they can say well actually I’ve allowed ten of my staff to go and actually, or allowed two of my staff to be train the trainers and I’ve had twenty of who’ve then gone through that, then it’s quite measurable, isn’t it?’* |
| Beliefs about capabilities | Confidence to deliver MECC training | Low confidence to deliver certain elements (e.g behaviour change theory, health inequalities, expectation effect) | AT3: *‘But it was just the models I was a bit unsure on, like I couldn’t, I think the COM-B I probably could reel off now, but at the time, like I said, it’s quite a lot of things get thrown at you and then to regurgitate them for someone else’*  AT11: *‘I think if you’re going to ask someone to then train that and they don’t have that background, if you make it too overly complicated, they’re going to go all right, yeah I’ll do the train the trainer and then they go nah, I don’t feel confident because I don’t actually understand that part myself.’* |
|  |  | Existing trainers are more confident in training delivery | AT3: *‘she’s very kind of open and confident and she is a trainer so I think she obviously wouldn’t struggle as much as I would’*  AT11: *‘I am probably pretty confident in doing training anyway. And then when I did the train the trainer it was stuff I knew about. I was like yeah, that’s fine right, I can do that no problem. So, I think my background really made it super easy for us to just transition. But as I say, I don’t think that’s as easy for other people if they hadn’t done training before and they haven’t come from a health background or a public health background, I think that could be a bit daunting’* |
|  |  | Professional confidence that you don’t need to be an expert | AT11: *‘All of the information’s there for you. It’s not, not rocket science. Just need to tell you about the information and the model and then off you go’* |
|  |  | Lack of confidence to deliver MECC training | AT8: *‘I think probably within the first couple of minutes of after doing it you think oh, I can do this, this is really good, but then as time goes on your confidence does fade, it wanes away the less you have to do with it’* |
|  | Perceived need and ability | Belief that staff already deliver MECC | AT8: *‘during the training I thought oh right, this will be good, I’ll deliver it, I’ll separate the team and kind of deliver it twice and thought yeah, yeah, this will be good. And then as time went on and I kind of thought about it and watched the team and interactions and different things, because there’s some really experienced members within my team, and then I think I got much less confident and thought, you know, it’s like teaching your granny how to suck eggs, like this doesn’t feel right, it’s what they’re doing day in and day out. So, I think confidence dropped off quite quickly’* |
| Beliefs about consequences | Beliefs around the impact of MECC | Belief in the effectiveness of MECC | AT4: *‘I have chosen to do that myself as part of my own health inequalities role. So, I felt this was something that it’s an at-scale programme, so the more people we can reach and the more people we can deliver MECC training to, then the more people have an opportunistic conversation which might have brought them to make some changes’* |
|  |  | Believes in the value of staff receiving MECC training | NA4: *‘I think the important thing is I think there does need to be more of an awareness for it. I think something like this should be part of everybody’s induction, I suppose, like especially in public health or in local authorities… which may have possibly helped me more in my first 12, 15 months in this role as well to understand public health a lot better … somebody starting out in public health and more into this world like especially like the NHS and things like that, I think having it as part of like an induction package, just understanding and increasing awareness of MECC would be really useful’* |
|  | Beliefs around attitudes towards MECC | Increase in perceived importance of MECC | PVC: *‘The importance of it - ie the significant changes that can be made, starting from just a conversation’* |
|  | Perception of the TtT model as favourable for optimal implementation of MECC | TtT helps promote and embed MECC within the organisation | AT11: *‘I think that the blanket approach is quite often a ticky box approach of well all of our staff are trained in MECC. Lovely but have you moved that forward? Have you embedded that into policy, into working practices, into everyday activities. And I think train the trainer should be that person who’s going well hang on, you know, can we do things differently… and then that can disseminate and be the voice for actually we need to do this, or we need to change this. You do need that because if you’re trained in it and then there’s no champion there going well can we do this, can we do that, I sometimes think it just gets well right we’ve done that and it’s in a folder with everyone’s name on and nothing happens’* |
|  |  | TtT provides unique benefits: specific knowledge and different perspectives | PT1: *‘And they gave a total different spin on it to what I would do from a public health and they were very human and funny and you could just see how they made it their own’* |
| Emotion | Passion towards MECC | Passionate about and sees the value of MECC | AT3: *‘I think so yeah, you have to kind of buy into what you’re selling really. But yeah, like I said, I kind of I understand it more because I’ve done like a few sessions. I know the importance of having a conversation…We have been converted. At first I was just thinking oh what’s this’* |
|  | Emotions around training delivery | Enjoys training delivery | NA4: *‘I quite enjoy sharing knowledge. I quite enjoy sharing knowledge more than anything and just talking to people about things. And I think it usually goes well but I’m quite a personal person, quite approachable as well. So, I think that kind of puts me in the right track for being able to share and train in the future’* |
|  |  | Fear of training delivery | AT9: *‘I felt still, and still do feel a little nervous beforehand, delivering the training so I can understand why people who aren’t trainers anyway, because we had in our group a lot of people from different organisations who hope to roll it out to their companies and it probably hasn’t happened. But one they weren’t trainers because they were really a bit nervous on the course like oh but what if I can’t answer the questions, you know. The sort of things you do worry about even as a trainer, and they weren’t trainers, so I bet that puts people off a little bit’* |
|  |  | Worries about technical difficulties and IT issues | AT9: *‘f you go to venues there’s always the IT issue. That’s always worrying for any trainer. I do quite a lot in the city hall and I’m sort of getting used to their IT and just plugging things in. I do a lot in community halls as well, like organisations similar to mine, and that’s when I’d have to take probably a projector because not all of them have a projector. But yeah, I’m fine going to the venues. It’s just you worry about the IT issues. But other than that, once you’ve got the IT up and running you know you’re just ready to go’* |
|  |  | Stress and discomfort around delivering MECC stats and theory | AT6: *‘I froze because I looked at the slides and went, I don’t understand them. I don’t get them. And when I looked and I had that conversation with my boss and I said look, I don’t get it and I’m struggling with it and it’s causing me stress. And she, because she’s from a higher level, she was like I don’t understand why, this is what you work with all the time. I says yeah, but these are heavy stats. These are Band 6. There’s this more experience talking about the stats, so if there’s any questions you can answer them comfortably. So, I avoided doing that. I don’t think I was very popular. But it was causing me stress. It was causing me a bit of wellbeing, mental wellbeing issues’* |
| Environmental context and resources | Top-down drivers to support cascade | No top-down support | AT6: *‘Management don’t have time, especially in a clinical setting, for you to go and say can I have four hours of your team, or three hours of your team, or even an hour of your team to talk about MECC. They do not have time. They do not like it’* |
|  |  | Top-down support for MECC model | AT5: *‘I think as the years have gone by there’s a lot, as I say, there’s a lot more kind of publications and things that have come out about MECC. You can see the kind of drivers, the links with government papers and things and all the sort of Public Health England stuff that they produced on MECC’*  AT7: *‘it’s embedded throughout, to be honest. I mean, I think we’ve got lots of different strategies and policies where MECC is featured within that... in terms of kind of the managers and leaders, absolutely get the whole concept of MECC’*  AT7: *‘the family hubs have really kind of picked it up and worked with it. They had management who already had had experience of MECC in the trust, so I think that definitely helped. That was steered at a very kind of leadership kind of level. Maybe, yeah, maybes leadership is something to do with it to help to kind of develop that vision, get it kind of, you know, developed and kind of cascaded in that way’* |
|  |  | Agreement or behavioural contract with manager pre-TtT | AT6: ‘*whoever goes on the training needs to have had a discussion with the manager on how they’re going to deliver it before they go on the training. And that maybe has to be checked on the day since everybody got their first MECC plan... And you only get on if you’re management understand that that is what an expectation is for your first delivery... So, I think it’s that, if you want to do that you need to have pre-permission from your manager to deliver it to a certain level and this will be done within the three months’* |
|  | Bottom-up drivers to support cascade | Bottom-up drivers | AT6: ‘you have to embed your awareness training first. It has to be there. Then those people that love MECC and then become champions will want to go to Train the Trainer.  To then be able to influence or support their clinical support, and when they leave, hopefully they’ve done enough to embed for somebody else. But if you get your new starts always flooding through, you’ll never have a shortage of champions’  NA5: *‘So, we’re busy transitioning and writing a new strategy for the organisation, which is going through the internal channels at the moment, and we’re hoping that we’ll be able to get MECC included in that new strategy. But obviously that needs sign off from the exec team. But we’re very keen to use that at the organisation’* |
|  |  | Organisational culture around MECC | AT7: *‘the different services within the council, they’re really on board with MECC. I think they really understand the role, it helps them to see even if their role isn’t around health and wellbeing, it helps them to see, actually how can I turn some of these conversation around that I have day in day out and shape it around health and wellbeing, bringing stuff in. So, I think people really do get it... I mean I think as an organisation MECC is very much a part of it’* |
|  |  | Cascade of informal training encourages staff buy-in and organisational culture change | AT7: *‘and then within the team having that conversation of right, so where do we have our MECC conversations, what points of contact do we have, what types of topics of conversation do we have, and how do we form a MECC three As framework around that, how do we ask them, how do we assist them, how do we act upon that, who are our referral signposts... you have that kind of it’s like an activity that you do rather than delivering, cascading training. Maybes it’s kind of having that conversation with the team to bring them on board, help them to understand where MECC fits, but then right, well how do we do this together as a team. So, maybes it’s more around rather than cascading training, maybes it’s around equipping people to having facilitated conversations’* |
|  | Useful resources with opportunity to tailor | Resources are provided | AT1: *‘I think it’s more motivating for people when they know that actually the work’s been done. We’ve provided everything for you, all we’re asking you to do is go out and deliver it. So, you don’t have to do any of the PowerPoints, you don’t have to source any of the resources, you don’t have to think of your own scenarios for your role plays because everything’s there. So, that really helps, and I think that will motivate a lot of people knowing that actually you’ve done a bulk of this for us. We’ve just got to go out and deliver it.’* |
|  |  | Opportunity to tailor resources to audience | AT10: *‘I liked the fact that they had different slides that you could sort of cut and paste in.... And the example that they gave during our training was health and wellbeing and I thought there’s no way that you’re going to get a (name of role) to go out and talk about diet, but they had ones about exercise and alcohol and stuff and I thought I’ll skim through those, those will be a bit more relevant, and I’ll use those as an example’* |
|  |  | Desire for physical resources | AT6: *‘a MECC pack would have been really good. If you go train the trainer, you should have picked up a bag with a whole load of stuff. You could go home, and you could read through and almost refresh yourself, not go on a computer screen and try and find stuff. Just the basic, the basic MECC pack. With stuff that you could then go to your first training session and feel quite comfortable in it’* |
|  |  | Guidance needed for accessing the relevant resources | AT4: *‘it’s which other of those hundreds of things do you need to include in your training sessions. So, I think a how to guide would really help. A step-by-step this is what you need to do to get your first session going and also when you go through the slide deck as I have been you have to obviously populate it with some of your own local information. So, I think a little bit of information in that how to guide are what you need to find to put in the presentation just to make it clear’* |
|  | Engaging and mobilising the right individuals in the right way | Expectations for cascade were not clear at sign up stage | AT6: *‘I think it’s the biggest waste of time I’ve ever been at because I didn’t actually understand what they were trying to deliver. So, train the trainer, to me, doesn’t mean anything. I’m trying to train the trainer. Who’s training what? Who’s training the trainer? I don’t understand that whole MECC train the trainer. It’s like if you’re a MECC trainer then you’re responsible so become a MECC trainer, reword it, do something but yeah and then we arrived, in all honestly, arrived at the course and everybody was in the same boat. They listened. They didn’t have a clue what they were trying to deliver and then they were told the expectation was they needed to deliver that four sessions in a year and nobody had a clue about it. So, that’s in all honestly why you lost or why our trust, lost a whole people because they weren’t prepared to do the training because they didn’t understand they had to deliver training. There’s not one person sat down in that first thing and went I’m here because I want to deliver the training for MECC and be an ambassador for MECC in my department. All they thought they were going to say is I’m at a course to become more aware of MECC so when I’m in my job I can be comfortable having MECC conversations’* |
|  |  | Only specific individuals have access to TtT | AT6: *‘so Train the Trainer model should only be delivered to people that are really ambitious and aware and want to deliver MECC to other people to sing from the rooftop saying this is MECC and we can come to your service, and we can make a difference’* |
|  |  | Reframing of MECC training to acknowledge staff may already be conducting MECC | AT8: *‘I would have to acknowledge their expertise and acknowledge that they’re doing this day in and day out. I would be very open and say the one thing I took away from the training is using it more with my colleagues, you know, with my peers, even I guess with family and friends, not just patients. You know, using this, that is one thing I certainly took away from the trainingl’* |
|  | Infrastructure to support cascade at organisation | Logistics of arranging MECC training | AT1: *‘a lot of the staff that we’ll be cascading this out to work shifts, a lot of them are part time or casual staff, and that they’re based across like five different centres. So, it’s logistically it’s been a bit of a challenge to get it all together’* |
|  |  | Designated time to deliver training | AT12: *‘we have a weekly CPD hour. So (every week) we do training. Not every single week, but it’s just called bite sized training, so we get people to come and do, deliver training, about all sorts of things we’ve had... So, it’s all beneficial stuff to us in that way, but then also things like the MECC training which are relevant but not in the same way, sort of thing. So, we delivered it within that slot… So that’s how it worked, but that’s a rolling programme, so we’ll probably put it on again next year and I’ll probably just do it next year’* |
|  |  | Embed MECC into existing programmes | AT13: *‘some of it might have to happen in trust induction or on a pre-existing training programme, like we've got preceptorship and care certificate, health and well-being champions is another one that we've got and we weave MECC into those. So they're quite good because we've got a bit of a captive audience and quite high numbers’* |
|  | Limited time | No time to deliver or attend MECC training | AT6: *‘We don’t have time for that. It’s hard enough for them to get the mandatory training done, you know, ESR. If you’re in an office job or luckily with public care, they’ll give that time, but if you got ward staff, they don’t even have time to do ESR. They’re miles behind ESR. So, how are you going to have that impact and say can you just stop for a minute and come along... It’s only three hours or two hours’* |
|  |  | Staff capacity | AT5: *‘she kind of just said it’s not something that we’ve got capacity to really put in place at the moment. We don’t want to bombard staff with other training when we’ve already got the smoke free, and we’ve got the alcohol training. So, it was less…seen as less of a priority back then’* |
|  |  | Competing priorities/ personal extenuating circumstances | AT8: *‘at that stage my motivation was quite high… within kind of 24 hours that all vanished because this is life, isn’t it? It throws these curve balls at you that you’re not expecting’* |
|  |  | Decision to shorten MECC training | AT5: *‘when we had conversations about it staff were just saying can we do it in two hours, can we do it in an hour. It would be much easier to release staff and I suppose the kind of weighing up whether it would be better to have people come on the training at all, or people not potentially come on and have this more detailed option. I think we’ve kind of decided that in the grand scheme of things it would be better for people to have some of that knowledge, a lot of people to have some of that knowledge and only a few people to have all of that knowledge’* |
|  |  | No time to attend support network meetings | AT6: *‘I don’t have time to attend that Teams meeting that’s really nice to discuss these things because I just don’t have time to book my time out’* |
|  |  | No time to deliver MECC training | AT8: *‘It’s hard really because I think they’ve tried to put the strategies in, in terms of supporting things. I don’t know, maybes it’s managers need to understand more that time needs to be given to the trainers to cascade the information. It’s really tricky. I think it all comes down to time and workload’* |
|  | Balancing utility with feasibility | Limited ability to lengthen training | PT2: *‘I don’t kind of go into the detail of the slide and how they might be using, how they might deliver them. Because whatever you do in terms of expanding that, it’s going to increase your timescale’* |
|  |  | Face to face training as preferred (balance opportunities for interaction and shared learning with logistics and reach of training) | AT8: *‘I think there’s pros and cons for each. I think obviously when you do face to face and it was a half day you’ve got to get there, you’ve got to travel home etc. so you’ve got that aspect of it. Certainly that day, as far as I know, everybody who was down to do the training turned up because the room was full, we had big tables and each table had several people on. So, I get Teams saves a lot of time and maybes you do get a higher percentage of people turning up because it is on Teams, it’s the click of a button so I can see pros and cons for both, but me personally, I learn much better in a classroom environment and networking with colleagues. I think that is you can’t put a price on that. It’s so valuable’* |
|  | Accessibility considerations | Difficulty in accessing resources | AT13: *‘..and make sure that people know how they're using the Futures platform. And, because it is complicated, you go on there, I don't know if you've used it? ..but you kind of go on and you’re like oh my god I cant get anything to work, I can't find anything on there and you know, there's always updates of slides and you know, this video, that video. So I do think people find it overwhelming’* |
|  |  | Accessible training | AT11: *‘So, I accessed that really easily and it’s on our training plan so anyone can access it, the core training’* |
|  |  | Online training gives the potential of a larger audience | AT5: *‘So, we’re delivering tomorrow. I’m delivering to about 200 preceptees and I suppose the benefit of online training is that you can do that. Obviously, we’d never have a venue that was available to deliver to 200 people, but they come on and do their inductions’* |
| Goals | Goals that facilitate cascade of MECC training | MECC aligns with other goals | AT10: *‘And it really summed up what we were doing… we were dealing with people who wouldn’t talk to anybody else and the MECC skills really encompassed what we were trying to do and we were trying to gather as much information in that 15 minutes that we could then refer on to other agencies and we could be that link to get someone support because they trusted us, so it really fits in really nicely with what we were trying to achieve’* |
|  | Goals (or lack of) to attend TtT and deliver MECC training | Lack of communication at sign up stage means that some attendees don’t want to cascade MECC (only attend MECC training) | AT4: *‘I think there needs to be two options for people for those who want to just do MECC so that they can use it personally. And I think because that wasn’t there, there were potentially people on the course who would use it in their own practice but maybe didn’t necessarily want to go and cascade it to others. So, I think there needs to be that differentiation. I know the point is to deliver it at scale. It’s that cascade but I do think not everybody would want or feel confident to do that’* |
|  |  | Aims to embed MECC training into existing processes | AT11: *‘My plan is I’m looking at right where are our services. So, implementation plan, we’ve got it onto (name of platform) which means anyone who has a council email address can access MECC. I would love to make it mandatory but that’s like not happening. So, it’s there so anyone can access it and it’s being advertised on (name of platform). We really need to get it onto sort of the newsletter or something like that, something that goes out to people’* |
| Intentions | Motivation and interest in cascading MECC training | Motivated to deliver MECC training | PSC: *‘this made me enthusiastic about how I can use this to support others moving forward’*  AT8: *‘I think at that stage my motivation was quite high and really our team morale was quite low, and I did think oh this could be a good thing to help kind of boost team morale’* |
|  |  | Individual wants to do the TtT training | AT2: *‘I take quite an open mind and I just go into it. I go into a learning situation because I want to, you know, and I’ve always been like that’* |
|  |  | Self-selected to attend TtT shows intrinsic motivation, mandatory training means disengagement | PT1: *‘but I think the people who sign up, if they’ve signed up voluntarily then it tends to happen. When they’ve been told, like in housing or adult social care you need to attend this because we want everybody on board, it’s a harder sell and you’ve got those people who throughout it, you know, you can just see straight away. So it’s maybe better self-elected to do the train the trainer’* |
|  | Planning and preparation | Planning where and how to deliver MECC training | AT7: *‘I don’t know if there could be an opportunity within that training session for a person to kind of sit down with like I suppose it’s like an implementation plan outline to kind of work through right, this is what I’ve learnt, this is what I’m going to do, so that it’s almost like a little bit of a plan of action of right, so I’ve just done that training and before I leave the room today this is what I’m going to do’* |
|  |  | Practice | PSC: *‘I don't think I need support, I just need to practice’* |
|  | Readiness before attending TtT | Planning where and how to deliver MECC training before attending TtT | AT6: *‘if you have somebody with a plan coming out and saying it’s already been discussed, this is where we’re going to go with it, and I want to deliver this, this and this and I’ve actually already emailed people. That’s what I would have said is the best way. So, the strategy afterward, they have to be prepared to be ready to go’* |
| Knowledge | Knowledge of the training content needed | Existing knowledge of MECC (e.g previous experience of MECC training) | AT3: *‘I know a couple of people just went straight to the Train the Trainer so they might have struggled a little bit more with not having that prior knowledge. Because it’s just a lot to take in, all the slides and things’* |
|  |  | Statistics and theory too high level | AT6: *‘It’s not rocket science. But when I sat down at that it felt like rocket science’* |
|  |  | No existing knowledge of MECC or associated content | AT6: *‘when you come to training and you leave, you should be comfortable about having that conversation, not panicking that you’re going to go and have a conversation with your peers about training them in MECC if you don’t understand what MECC is’* |
|  |  | Familiarisation with the course content | PSC: *‘I just need to familiarise myself with the content and practice it’* |
|  |  | Knowledge increases with repetition | AT12: *‘yeah it was a bit daunting at first because I’ve never done it before, but it’s like anything, isn’t it.. the more you do it the more familiar you are with everything’* |
|  | Knowledge associated with the TtT model needed | Atendee did not know they were attending a TtT course | PSC: *‘I was not aware i was attending train the trainer training. I thought i was attending MECC training’* |
| Memory, attention, and decision processes | Learning process during TtT training | MECC training immediately followed by TtT as overwhelming | AT1: ‘I think what didn’t work on that day for some of the team was the Train the Trainer directly following, like immediately following the core training. I don’t think that really worked because as I say there were a number of people in that core MECC training who had no experience with MECC, never heard of MECC, didn’t know what it was, who went through the core training and came out the other side a little bit sort of confused and bamboozled and then within 30 minutes later they’re then going into Train the Trainer. And I just think that was too much for them on that day’ |
|  |  | No framing of MECC before policy and theory | AT13: *‘it felt like we were all sat in the training for a good hour or so before MECC was even mentioned. So it was, and other colleagues did say that so what we did was with, we obviously have kept in health inequalities, etc. But we moved to the MECC information a lot earlier on in the training. Just so that people werent getting to sort of an hour plus and thinking, well, this is all really interesting, but where does the making every contact count element come in?’* |
|  |  | TtT training catered to a range of learning styles | AT1: *‘they tried to use as many different approaches as possible as in it wasn’t all just somebody stood at the front talking to the group. You know, there was some things in there, group activities, resources put out on the table for smaller groups to work in. Larger group flip chart brainstorming sessions, if you like, so there was a good variety in the thing that they used in the training and the styles that they usedl’* |
|  | Retaining memory after TtT training | Time delay after receipt of TtT training means content is forgotten | AT5: *‘Part of me thinks that a lot of, because I always think well why on earth are people coming to do a Train the Trainer session and then not delivering it, because even I understand the people that have done the training a couple of years ago and then COVID’s you know, life has changed dramatically since then. Easy to forget. It’s easy to forget a training session if you haven’t delivered it for a few weeks, never mind a couple of years. So, I get that. So, it almost can write out the ones that did it a few years ago’* |
| Optimism | Optimistic attitude towards MECC training delivery | Positive and assured around training delivery | AT10: *‘I didn’t necessarily feel like I needed formal slides. If I went through how to have the conversation, I could just say give them a couple of examples and match up with the questionnaire and then use it that way, so I was fairly happy that they’d given us what we needed… So, I was fairly happy just to get out and get going with it’* |
| Reinforcement | Intrinsic or extrinsic consequents for the individual | No reward for delivery of MECC training | AT6: *‘you ask me to go and do some extra training to go and deliver to Band 5s, Band 6s, Band 7s so they can make a difference in a clinical environment. So, I’m teaching people that are higher paid than me, I’m delivering an impacted thing so they can become better so they possibly can climb the ladder more because they’re MECC trained, and they’ve become more aware. So, actually that’s probably quite a big thing in their career pathway, but I get nothing for it. Tell me where that expectation is fair’* |
|  |  | Incentives | NA1: *‘And I suppose if someone’s going to say well is it worth my while being one of these trainers, it’s the sell, like what is it that you will then be able to go and do and influence and how good will that impact be for you personally and for those that you’re training thereafter. And will you be able to see a benefit somewhere down the line. Or an impact that you’ve had’* |
|  |  | No feedback when delivering online | AT12: *‘that stony silence when nobody would answer me it was like oh God, no. And I was thinking, I said everybody disappeared, they were all gone and has everybody slipped off because it was so boring’* |
|  | Extrinsic consequents for the organisation | Reward for organisation | AT13: *‘So we have that organizational view of who's done the training rather than it sitting on regional app and then not, it sounds awful but that’s not doing us any favors from an organizational point of view, what we need it on is like the electronic staff record’* |
| Skills | How to be a MECC trainer | Existing skills associated with training delivery | AT1: *‘every week that I’m standing up delivering health lifestyle sessions and stuff like that and using the PowerPoints and stuff, so in that sense absolutely fine, it’s not something that’s completely new. So, and a lot of the team are the same, to be honest. A lot of the team have to do very similar things with the programmes that they deliver on, so I think we’re all pretty confident about it’* |
|  |  | Ability to tailor MECC training whilst retaining quality and consistency | AT10: *‘you’ve got to have the same information, so the slides that we tweaked in the past we used to go out into the community and deliver (role specific) advice. The advice on the slides had to be the same no matter who was delivering, but you might have different transitions, you might have different pictures or different colours, you might, you can tweak the graphics and tweak how you’re going to explain something without changing what you’re actually explaining, but make sure you’re getting all the same content. But you can do that in different ways’* |
|  |  | Advice on how to be a MECC trainer | AT8: *‘So, (principal trainer) gave all the tips of the trade and, you know, of what, you give examples of what went bad for him when he was presenting, what went good for him. So, they did very much give you those, try to give you those tips so you could develop the skills to be a good teacher’* |
|  |  | MECC TtT insufficient for non-trainers | PT2: *‘I wonder if part of that problem is you’re getting people who are being sent on this course who’ve got no experience of delivering training or talking in front of people and then they’ve got three and a half hours with us and then they’re expected to go and deliver training’* |
|  |  | Non-trainers need the section on the ‘how to’ of training delivery | AT5: ‘*I think the Train the Trainer bit on the end was really useful because I think people aren’t necessarily natural trainers. They haven’t necessarily done a lot of training before, so I think you do need that kind of how to, almost, at the end of it. You can’t just receive the content and then go – or you can’t necessarily just receive the content and go away and just do it... especially if you haven’t done any training ever or for a long time’* |
|  |  | Training acts as a refresher for existing trainers | PSC: *‘it was an excellent refresher and offered new skills and techniques to include in my current training’* |
|  |  | Staff in healthcare settings don’t need MECC training | AT6: *‘To be quite honest with you, I think it was like we maybe don’t understand what our staff are already doing so you’re trying to label something they’re already doing... I don’t think they need training’* |
|  |  | Health/NHS staff more used to *see one, do one, teach one* model of learning new skills | AT11: *‘the trainers are NHS background, so by default I’m an NHS learner, sort of kind of we’ve all been, so regardless of your learning style, we’re probably conditioned to learn in a certain way. Well, it is because NHS background’s usually we’ll train you, see one, do one, off you go kind of background. And I think that’s what the training is like, whereas my colleague who did the training with me is then coming with me to do further, you know, to observe the training that I do, and I wonder if that’s because she’s not that kind of see one, do on, off you go kind of learner’* |
|  | Interactive TtT training needed | No opportunity to practice training delivery | AT1: *‘I was kind of expecting a little bit of you need to be able to go away from today feeling comfortable training other people in MECC, so let’s practice delivering the training of MECC.. I guess I just thought I’m being trained to do something so will I get the opportunity to practice doing what I’m being trained to do. So yeah, I think just for me, I just thought there might have been an element of let’s practice some delivery’* |
| Social influences | Peer support as essential to facilitating cascade | Individual facilitates peer support | PT1: *‘we then would say OK then, (name of location) we’ve got nine trainers and they’ve got two more coming on, anybody going to deliver so the new trainers can actually sit in or shadow or co-deliver, and so they’ve got like a lovely group now that basically talk to each other and say well can you cover that one’* |
|  |  | ‘Buddy up’ delivery | PSC: *‘Potentially someone to deliver it with me, to help me know I am doing it right’* |
|  |  | Support from other staff at organisation | AT12: *‘it was good to have (name), who’s our sort of link worker for that, because she gave me loads of tips and everything.. so that was quite good and then she just gave me some good tips about things to do and ways to be, so that was great. Had I not had her, it would have been probably a different experience, and that’s helped me sort of do training, sort of think about the training that I’m going to deliver’* |
|  |  | Support from those in a similar setting or location | PT1: *‘but also you need it as different sectors or abilities so again if you’ve got people who are delivering in schools you really need people who are delivering in schools to recognise what those parameters are rather than fit in with the (name of location) group because nobody else might be delivering at schools’* |
|  |  | Peer learning | AT8: *‘I would love to hear like the lady at (name of location) who was really enthusiastic but she had a lot of challenges in how she was going to deliver it and things, I think then, if you then got the chance to be in a peer support kind of group afterwards and say right, did you manage to do it, how did you overcome these obstacles, how did you have the time, etc. etc. I think that would be brilliant. It would be really good and quite motivational’* |
|  | Additional support after TtT training required to facilitate cascade | Continued support | PT1: ‘*if you put the right systems in place you can literally nurture people and join them together and send out that email to say hey, I notice you haven’t come into the monthly meeting for like the past 12 weeks, I’m just wondering if everything’s OK. And I know we now have a regional MECC steering group and to be able to bring people together, but it’s also that when you first go through the training you do need that follow up and you’ve got to have the time and capacity and buy in to do that’* |
|  |  | Opportunity to shadow/modelling after TtT | PT1: *‘It took a lot of work. In the beginning, if you can imagine, if you’re not a trainer but you’re saying come along, sit in. I mean the (name of organization) fellas probably sat in three sessions after the train the trainer and watched and took notes… They sort of really developed their confidence to then say right, I’ve got this session’* |
|  |  | Support required to access resources | AT2: *‘but again I think there should have been more help there and pushing you and saying yeah, you know, get on the app’* |
|  |  | Support needed after to retain accountability | AT2: *‘The actual teaching process, the actual course structure was good. As I say, I would put more emphasis on after. Say well what are these individuals doing now? Do they do anything as in teamwork or have they done anything together. So, again, it’s the follow up, isn’t it? The following up’* |
|  |  | Lack of guidance after TtT | AT7: *‘when a person leaves that session there’s still quite a lot of work to do to adapt it for them and I don’t know if that’s something that can be done in the training. Because obviously it’s very specific to the individual and the organisation. They almost like, I suppose yeah, so it’s not that I don’t like it, but I think maybe you do the training and then the person is leaving but it’s kind of well what are they going to do with that now.... it’s kind of maybe a bit too left to the individual to then take it forward.’* |
|  | Social dynamics with potential trainees | Communicate worries about presenting with potential trainees | AT8: *‘maybes I need to ask the team would you like this and I would be very open and honest with them and say this is going to be my first time presenting this, you know, so bear with us type thing’* |
|  | Social dynamics with TtT training providers | Utilise existing networks to advertise TtT training | AT9: *‘(name) in (name of location) Council advertised it, and I might have also got, because obviously I’ve links with the RSPH, they might have advertised it.’* |
| Social/professional role and identity | Congruence of MECC with social or professional identity | MECC as part of professional identity | AT1: *‘I just thought why not really. I’ve done so much of MECC in the past and I’d been trying to implement it into my working role for such a long time that, and a lot of the team that I’m working with haven’t had as much or if any experience with MECC, so I thought it was just only the right and natural and the fair thing to do for me to say I’ll do the Train the Trainer.’* |
|  |  | Identify MECC champions or advocates to complete MECC TtT | NA6: *‘our thoughts would be that we probablies getting some champions across the organisation, you know, people who are already keen to do it. And working within our health and wellbeing strategy at the moment we’ve got some key people that we can, that we know that they would be more than keen to actually undertake the training’* |
|  | Ownership of MECC training delivery as part of role | MECC trainer as part of role | AT7: *‘It was part of my job description as well, so I knew that that was this deliverable that had to kind of be delivered on. So, there was that kind of motivation in terms of actually this is my job, I’ve got to kind of cascade this. Which I suppose is a bit different to maybe some other people that are attending the Train the Trainer where they might not have that kind of either written into their job description’* |
|  |  | Opportunity to tailor gives sense of ownership over MECC training | PT1: *‘And they’d just made it their own and it was just like well that’s my job done. All it took for me was you’re the experts, I’m not. I’ll give you these slides, but it doesn’t mean anything unless you’ve got ownership’* |
|  | Identity of potential trainees | Resistance from staff to be MECC trained | AT6: *‘I’ve avoided it like the plague until…I didn’t avoid it deliberately, I did try and push it to start off with, then obviously when I was still in my old position it was just it wasn’t happening. It wasn’t happening. It was there was too much resistance so you just kind of hide away from it, and that would be, probably I would have said the majority of people that have tried to do the Train the Trainer’* |
